# Determining international spread of novel B.1.1.523 SARS-CoV-2 lineage

**DOI:** 10.1101/2021.11.21.21266655

**Authors:** Lukas Zemaitis, Gediminas Alzbutas, Dovydas Gecys, Andrey Komissarov, Arnoldas Pautienius, Rasa Ugenskiene, Marius Sukys, Vaiva Lesauskaite

## Abstract

Here we report the emergence of variant lineage B.1.1.523 that contains a set of mutations including 156_158del, E484K and S494P in Spike protein. E484K and S494P are known to significantly reduce SARS-CoV-2 neutralization by convalescent and vaccinee sera and are considered as mutations of concern. Lineage B.1.1.523 has presumably originated in Russian Federation and spread across European countries with the peak of transmission in April – May 2021. The B.1.1.523 lineage has now been reported from 27 countries.

## INTRODUCTION

The emergence of severe acute respiratory syndrome coronavirus 2 (SARS-CoV-2) in late 2019 led to the ongoing Coronavirus Disease 2019 (COVID-19), now a global pandemic with more than 230 million cases of infection and about 5 million deaths worldwide ^1^. On January 5, 2020, the first whole genome sequence of 2019-nCoV was completed by Wuhan Institute of Virology, China Centre for Disease Control and Shanghai Public Health Clinical Centre of Fudan University ^2^. From this point on, genome sequencing has played an important role in vaccine development, understanding viral evolution and epidemiological characteristics. In many countries, SARS-CoV-2 sequencing has been implemented at the national level as a tool for epidemiological management^3^.

A large dataset of SARS-CoV-2 genomes has been collected in the GISAID database, which now contains more than 3.7 million sequenced genomes from around the world ^4^. As of May 31, 2021, World Health Organisation (WHO) has proposed designations for global SARS-CoV-2 variants of concern (VOC) and variants of interest (VOI) to be used alongside scientific nomenclature in communications about variants to the public ^5^. This list includes the variants on the global list of WHO VOC and VOI and will be updated as the list of WHO changes. There are currently three SARS-CoV-2 VOCs: Beta, Gama and Delta. The variant previously classified as Alpha (B.1.1.7) has been reclassified as de-escalated due to the drastic decrease in prevalence in the EU/EEA ^6^.

Through the routine analysis of National Lithuanian sequencing results from national sequencing efforts coordinated by National Public Health Surveillance Laboratory, we have identified a novel SARS-CoV-2 variant classified as B.1 by PANGO but containing multiple S protein mutations associated with effects on immunity (https://github.com/cov-lineages/pango-designation/issues/69), such as E484K; 156_158del; S494P. Preliminary phylogenetic analysis indicated that this variant has a distinct viral lineage that may have originated in Russia. We reported this variant to the PANGO curators and gave it the new phylum name B.1.1.523 (yhttps://github.com/cov-lineages/pango-designation/issues/69). On 14-July-2021, WHO added this variant to the list of variants under Monitoring section. By using bioinformatics tools, we performed a detailed analysis of this lineage and disclosed our findings about this variant or the mutational subgroups typical of this variant.

With this report we aim to share a detailed analysis of discovered variant, evaluate its origin as well as predict potential epidemiological impact and risks.

## RESULTS

### Mutation review of B.1.1.523

Several mutations in S region have been observed in B.1.1.523 variant, from which 156_158del, E484K, and S494P are considered as an attribute for VOCs (Fig. 1). According to previously reported data, the 156_157del and G158R mutations in the Delta variant are matching to the same surface as the 144 and 241–243 deletions in the Alpha and Beta (B.1.351) variants, respectively. These altered residues are found in the NTD ‘supersite’ that is targeted by most anti-NTD neutralizing antibodies, thus providing a mode to dodge immune system ^7^. Moreover, E484K mutation also contributes to SARS-CoV-2 immune system evasion. Several recent studies have observed that E484K may significantly reduce convalescent serum neutralization ^8,9^. Additionally, it was observed that S494P mutation is related to 3-5-fold reduced SARS-CoV-2 neutralization in sera), however, this mutation was not as potent at neutralization as E484K ^8,10^. With a combination of 156_158del, E484K, and S494P mutations, B1.1.523 lineage should remain on epidemiologists watchlist as one of the most concerning SARS-CoV-2 lineages.

**Figure 1.**
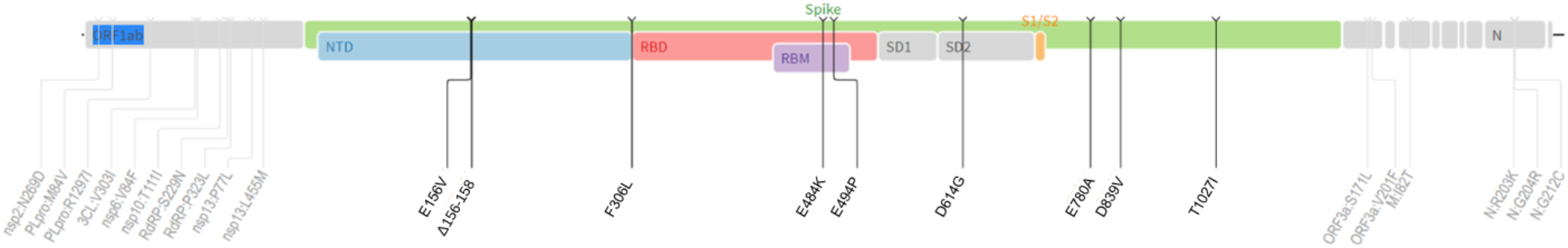
Mutation overview in B.1.1.523 lineage. Several other mutations have been observed in the spike-protein sequence of B1.1.523 variant, including E156V, F306L, D614G, E780A, D839V and T1027I.

In addition to 156_158del, E484K, and S494P more than 70% of genomes attributed as B.1.1.523 lineage possess F306L, D839V and T1027I in Spike and a set of substitutions in ORF1a (NSP3:M84V, R1297I; NSP2: N269D; NSP5: V303I; NSP6: V84F; NSP10:T111I), ORF1b (NSP12:S229N, P323L; NSP13: P77L), ORF3a (NS3: S171L, V201F), M:I82T, N:G212C (See Fig. X). T1027I substitution in Spike is common for VOC Gamma (P.1). Little or no information is available on these mutations.

### Origin and formation of key S protein formation of the lineage

One of the objectives of the analysis was to determine if the two clusters of mutations, responsible for immunity resistance, were acquired by sequential mutations or if this is a result of recombination events. Initially a list of data entries comprised of the sequences classifiable as B.1.1.523 lineage together with the closest sequences based on identity was used to construct a maximum likelihood (ML) tree. The size and overlap between the two data sets is depicted in Fig. 2.

**Figure 2.**
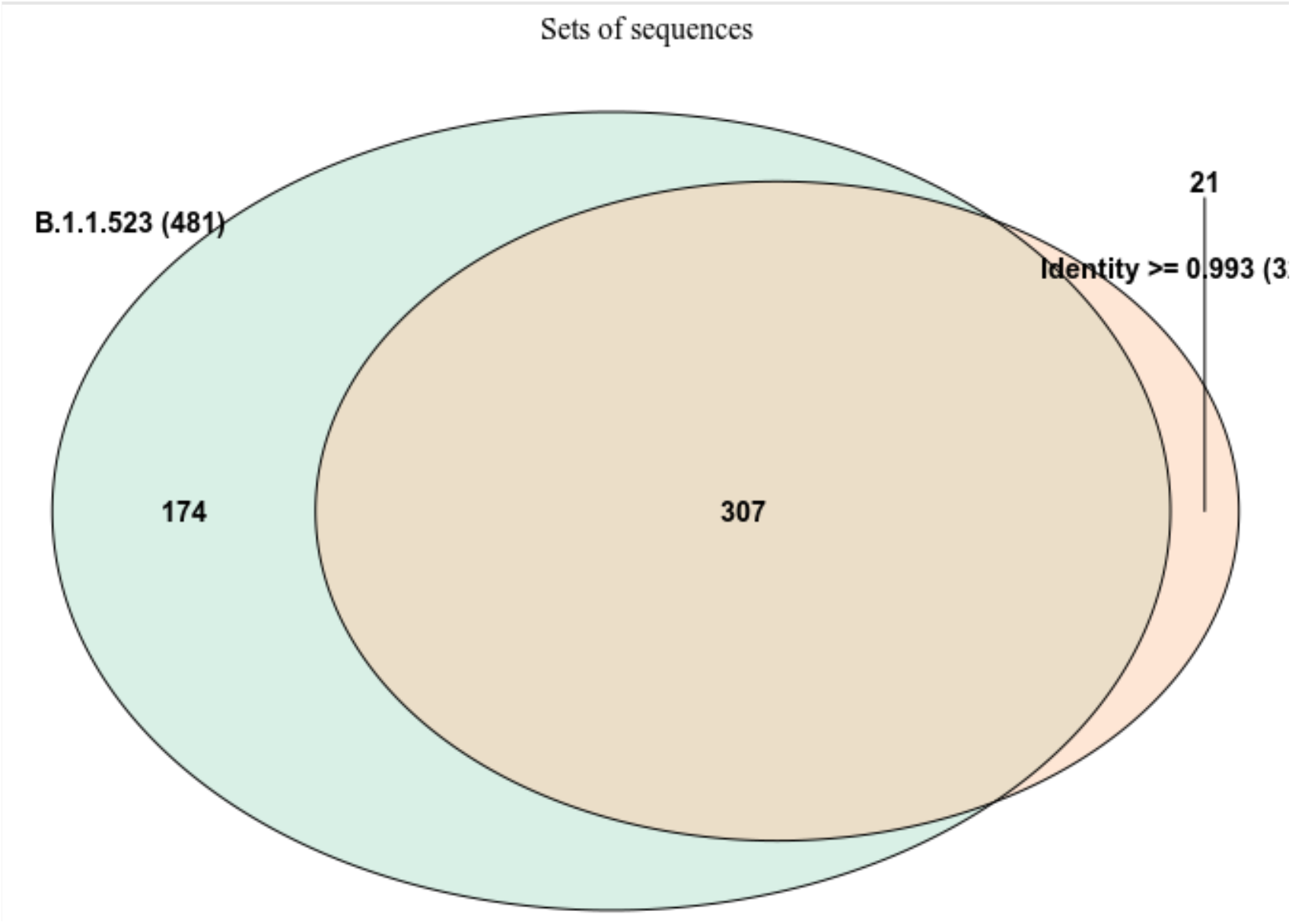
The overlap between the two data sets used for focused ML tree. The sequences were either chosen based on Pango assignment or by the identity with a B.1.1.523 lineage sequence (EPI_ISL_1590462). Most of the sequences, with high identity (> 0.993) to the Latvian B.1.1.523 lineage, were classified as belonging to the B.1.1.523. However, 21 (6%) sequences were not assigned to the B.1.1.523.

Generation of ML revealed several interesting properties of B.1.1.523 (Suppl. Fig. 1). At the base of the lineages leading to the B.1.1.523 sequences having a full set of expected S protein mutations branches away in clusters of sequences having the triple S:156_158del deletion. The sequences having the additional substitutions at S:484 and S:494 positions emerge further in the evolution. However, here, we have no clear indications of that the mutations S:E484K, S:S494P are introduced sequentially to form the B.1.1.523 lineage.

We have observed that some sequences which originated from progenitors with full set of expected substitutions at 484, 494, 156, 157, 158 S region positions, have underwent a reverse-type mutation to wild type variants. Such events are highly unlikely and usually are caused by erroneous sequence assembly or could be an indication of low-quality data. Additionally, in order to discern the cases where the mutations comprising the two regions of potential enhanced resistance to immune response have been combined phylogeny analysis of all S protein unique variants was performed. As it is depicted in Figure 3, there are at least three distinct cases where E484K and/or S494P have been combined with the 156_158del. Phylogeny analysis of S protein indicated that lineage assignments by Pango sometimes can be misleading. Furthermore, relying on plain assignments could hide novel developments of SARS-CoV-2. For example, some sequences from Turkey that were assigned as lineage B.1.1.523 originated from distinct clusters based on S sequence (Fig. 4A) and were evidently evolutionary distant from other B.1.1.523 lineage genomes.

**Figure 3.**
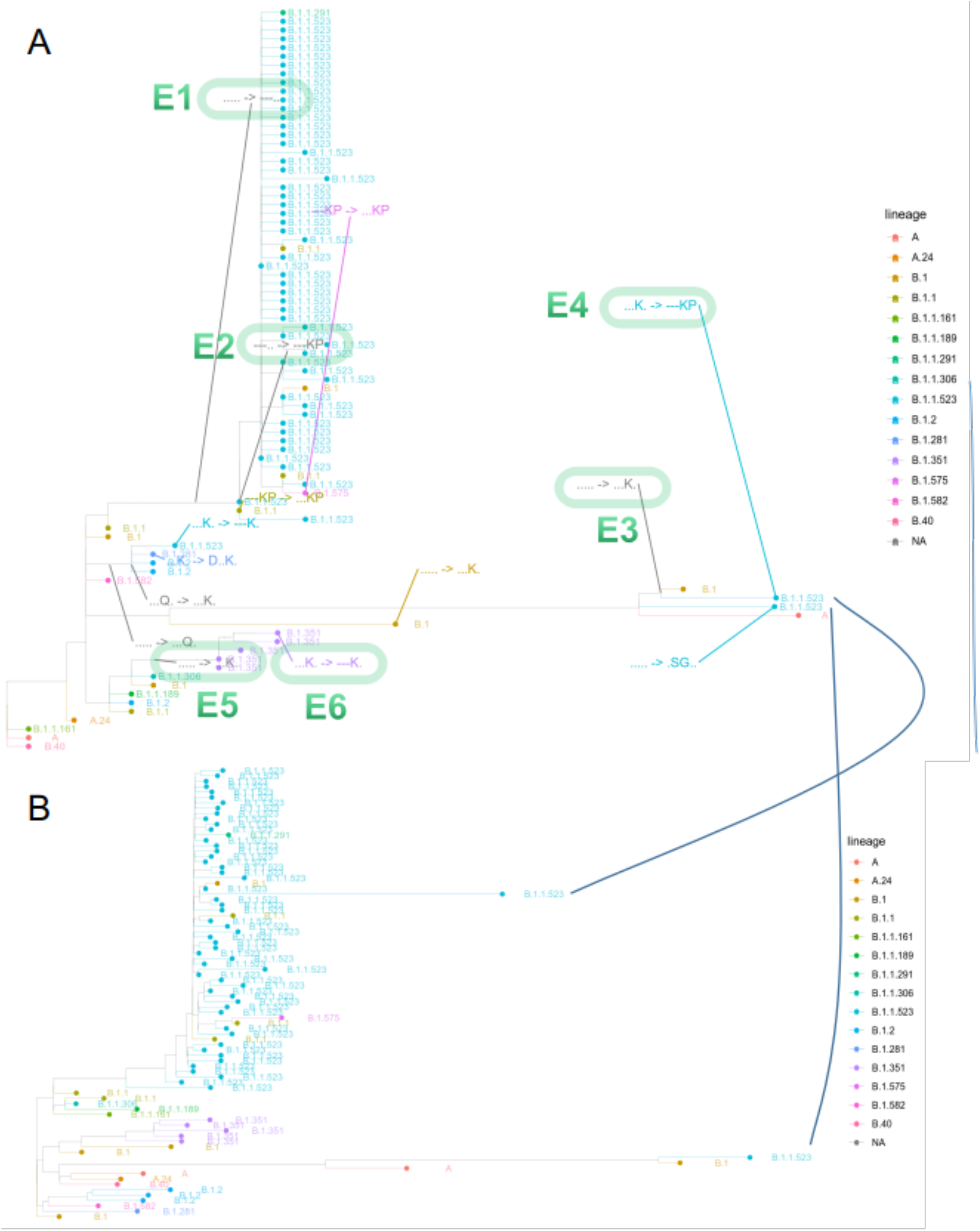
Phylogeny based on S protein sequence. (A) The tree represents a maximum likelihood tree based on all unique S protein sequences of the genomes deposited to the GISAID. The visible subset of the tree matches lineages that lead to branches having 156_158del and E484K or S494P mutations. The arrow “->“ indicates haplotype transitions detected by comparison of parental sequences with their offspring variants. The five-letter haplotype strings match 156, 157, 158, 484, 494 positions of the S protein with “.” meaning the wild type. The green ovals and their number indicate prominent transitions that are discussed in the main text. (B) The lower tree matches maximum likelihood tree based on whole genomes of the cases visualised in the upper tree. The black solid lines indicate a match of nodes for two B.1.1.523 sequences from Turkey in the phylogenies based on S protein and whole genome.

**Figure 4.**
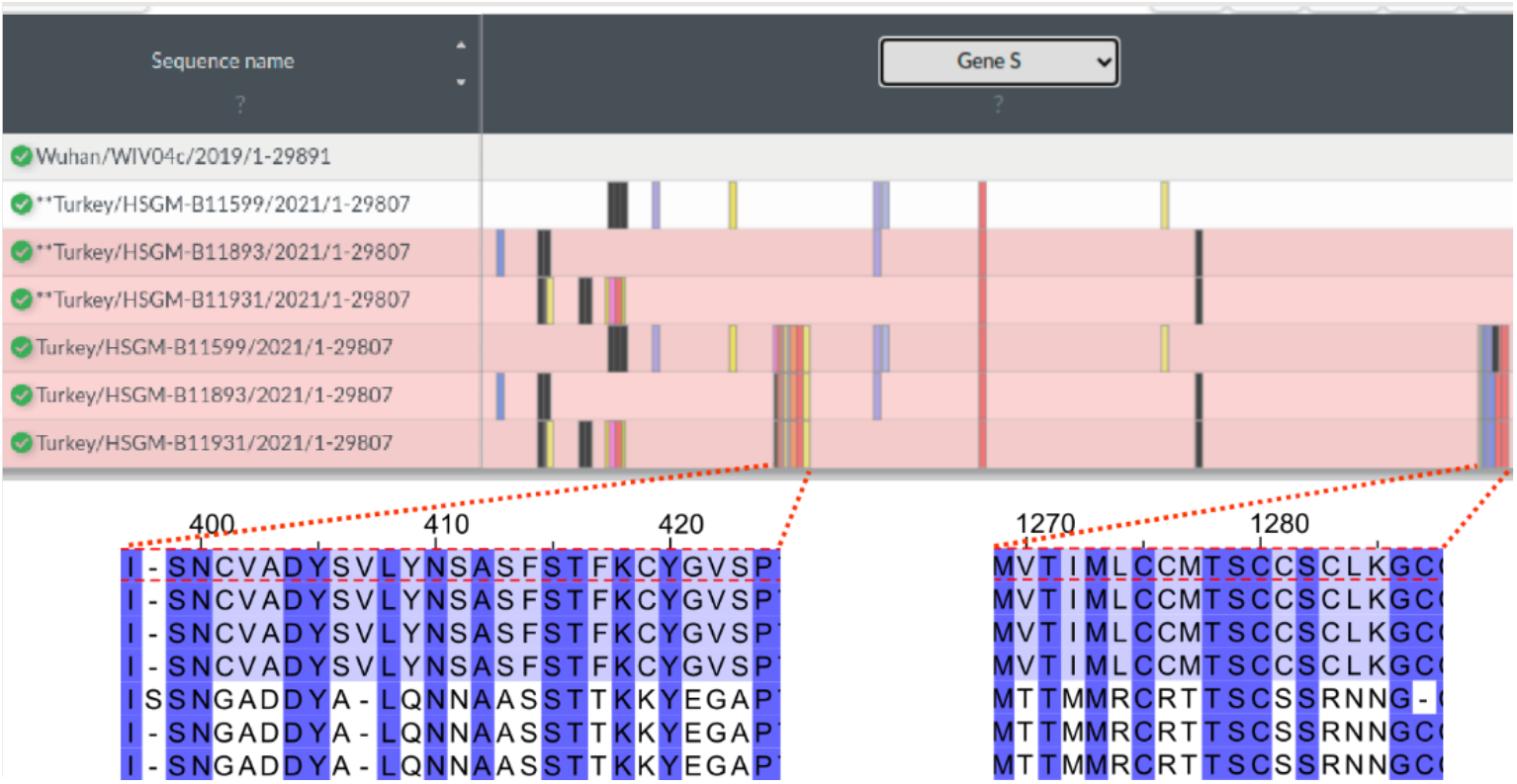
S protein region of B.1.1.523 sequences from Turkey. Turkey/HSGM-B11599/2021 and Turkey/HSGM-B11931/2021 were classified by Pangolin as belonging to B.1.1.523 lineage. The top figure is a snapshot from the Nextclade analysis. The top sequence is the reference sequence hCoV-19 that is used by GISAID. The asterisks ** indicate sequence from Turkey variants that have their two most variable sequences swapped with corresponding regions from the refence sequence. The two lower graphs indicate the sequence alignments from the two extremely variable fragments. The order of sequences is the same as in the upper graph: reference sequence, three Turkey sequences with swapped fragments, three original Turkey sequences.

Transitions E1 (introduction of the triple deletion) and E2 (introduction of E484, S494P) (Fig. 3) indicate a pathway where the majority of B.1.1.523 lineage sequences took. As in the case of data given in (Fig. 3) we do not detect sequential acquisition of E484K or S494P, and immediately next to the triple deletion we see the two aforementioned additional mutations. This could indicate a potential recombination event, but it is most probably due to lack of insufficient data.

An interesting case is with two highly diverged Turkey sequences that are classified by Pango as B.1.1.523 lineage. In this case following the most likely scenario at first the E484K was acquired and only then the S494P and E156_R158del were acquired (Fig. 4). As in the case discussed before, the sequential acquire steps are lacking from the data. Most probably the sequencing data was not comprehensive enough to reveal the full picture on how the combination was formed. In addition, these two highly diverged sequences indicate that there exist vast uncharted territories of COVID-19 evolution as we get only sparse sequencing data from Central Asia regions where the COVID-19 infection are barely controlled. An evident third case emergent combination of immune response hindering mutations from distinct S protein regions are highlighted by E5 and E6 at Figure 3A. In this case within the B.1.351 lineage at first the E494K mutation was introduced and then E156_R158del followed. The presented data evidently shows that the immune response hindering mutations from RBD and NTD domains have been combined in one protein at least three times. In two of them E484K occurred first, in one case the first one was the triple deletion. Most probably these are results of independent mutational events.

In Figure 5 the Turkey sequences are highly diverged from the wild type based on the region variation of S protein. Based on the GISAID blast searches (performed at 2021 10 1 07) the first variable region (bottom left, Fig. 5) from Turkey/HSGM-B11599/2021 was found in 7 sequences and the second variable region (bottom right, Fig. 5) was found in 4 sequences. The sequences have been deposited in the GISAID during several submissions. Other genomic regions of the Turkey sequences showed in the Fig. 4 do not contain such large SNPs clusters as the ones showed in the S protein. This indicates the changes in the placement of the three Turkey sequences upon switching the two extremely deviated fragments with corresponding fragments with reference sequence.

**Figure 5.**
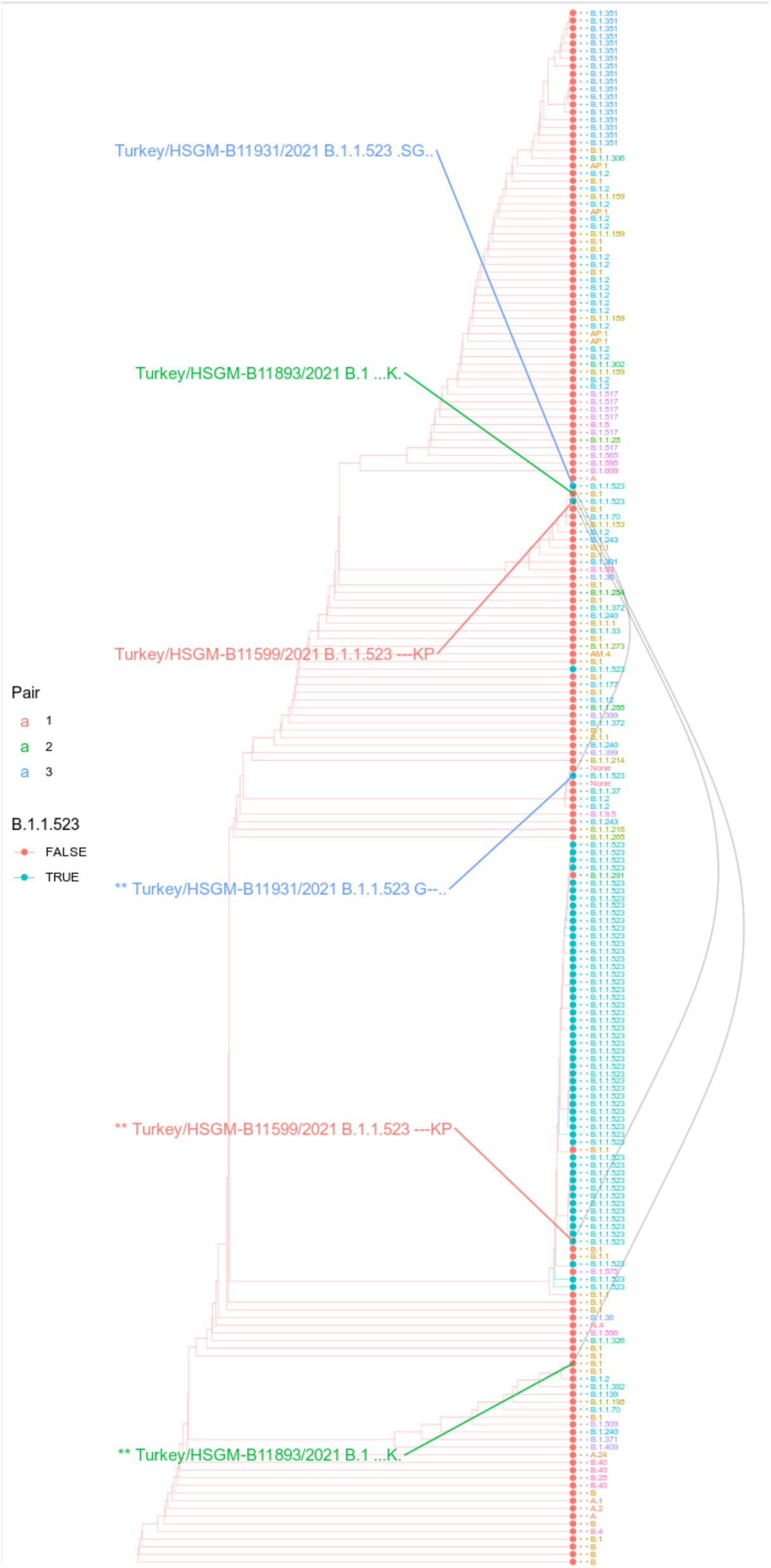
Phylogeny based on S protein sequence with modified sequences from Turkey. The cladogram of maximum likelihood tree that includes the three pairs of Turkey sequences with their variable regions either swapped with counterparts of reference sequences or being left original. Colour of the tips of leaves indicates if they are being classified as B.1.1.523. Tip labels indicates their Pangolin assignment with colours indicating different lineages. The different tips connecting grey lines indicates a Turkey sequence. The asterisks (“**”) in front of the sequence name labels indicates the sequence variant where the variable regions were swapped with corresponding regions from the reference sequences. Next to the sequence labels the haplotype at positions 156-158,484,494 with “.” indicating the wild type and “-” - a gap. A Turkey sequences.

### B.1.1.523 spread worldwide

Till August 31^st^, 2021, 459 B.1.1.523 sequences spread across 27 countries have been published in the GISAID. According to analysis results, B1.1.523 has originated in Russian Federation and spread across European countries (Fig 6). The sequenced clades peaked at week 25 and then subsided. In total, 95 transmission clusters have been identified. The peak of B.1.1.523 transmission intensity was around April - May 2021. The most numerous transmission clusters were detected for with MRCA’s originating from Germany and Russia.

**Figure 6.**
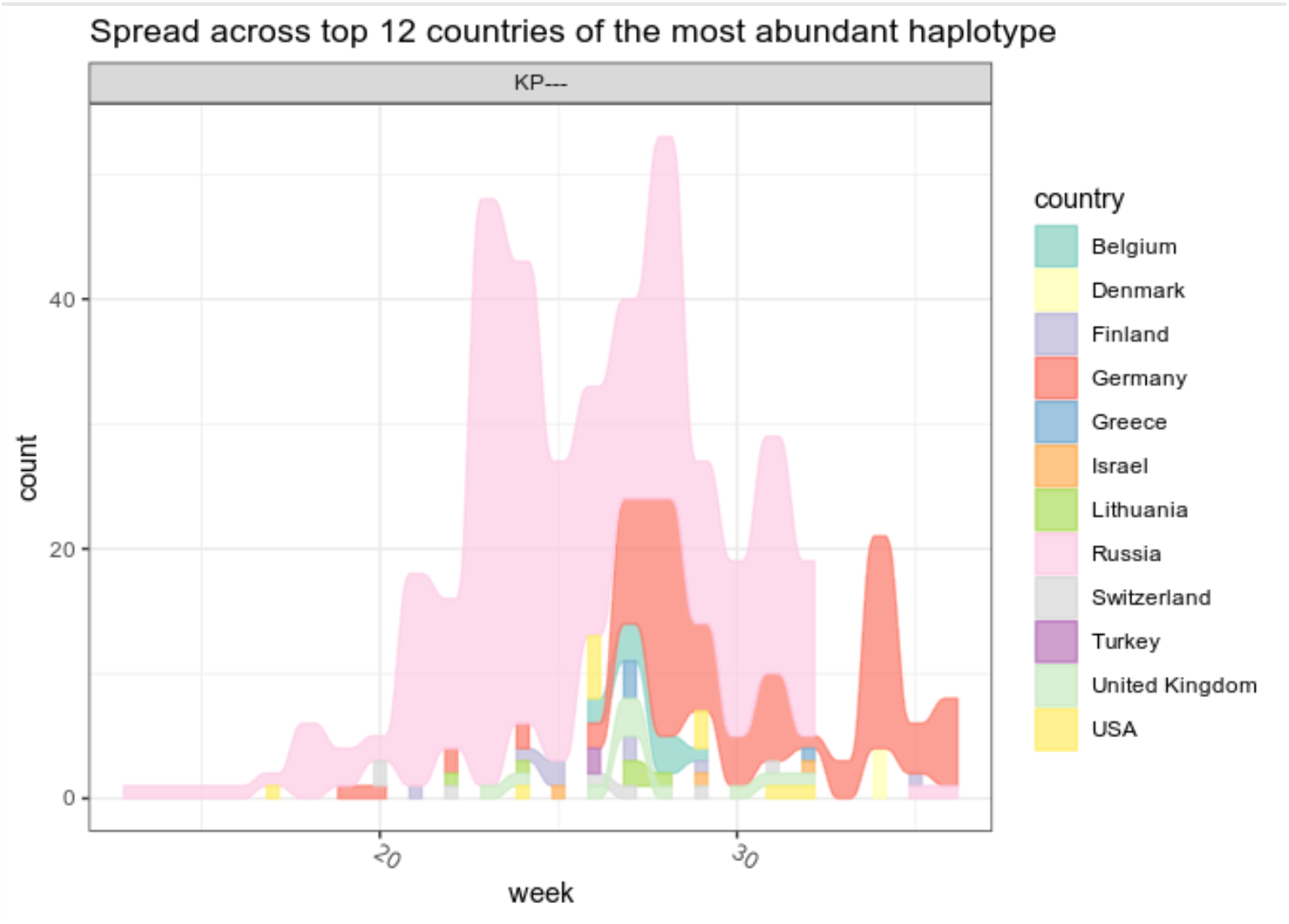
The distribution of cases of the lineage B.1.1.523 across countries at different time points. The “0” time point indicates the date of the earliest lineage sequence uploaded on the GISAID database. Only sequences that have the typical set of S mutations were considered (E484K, S494P, 156_158del). Only the cases which are corresponding to top 12 countries with the most abundant detection rate are included in the underlying data. The top 12 countries correspond to the 93 % of all cases.

**Figure 7.**
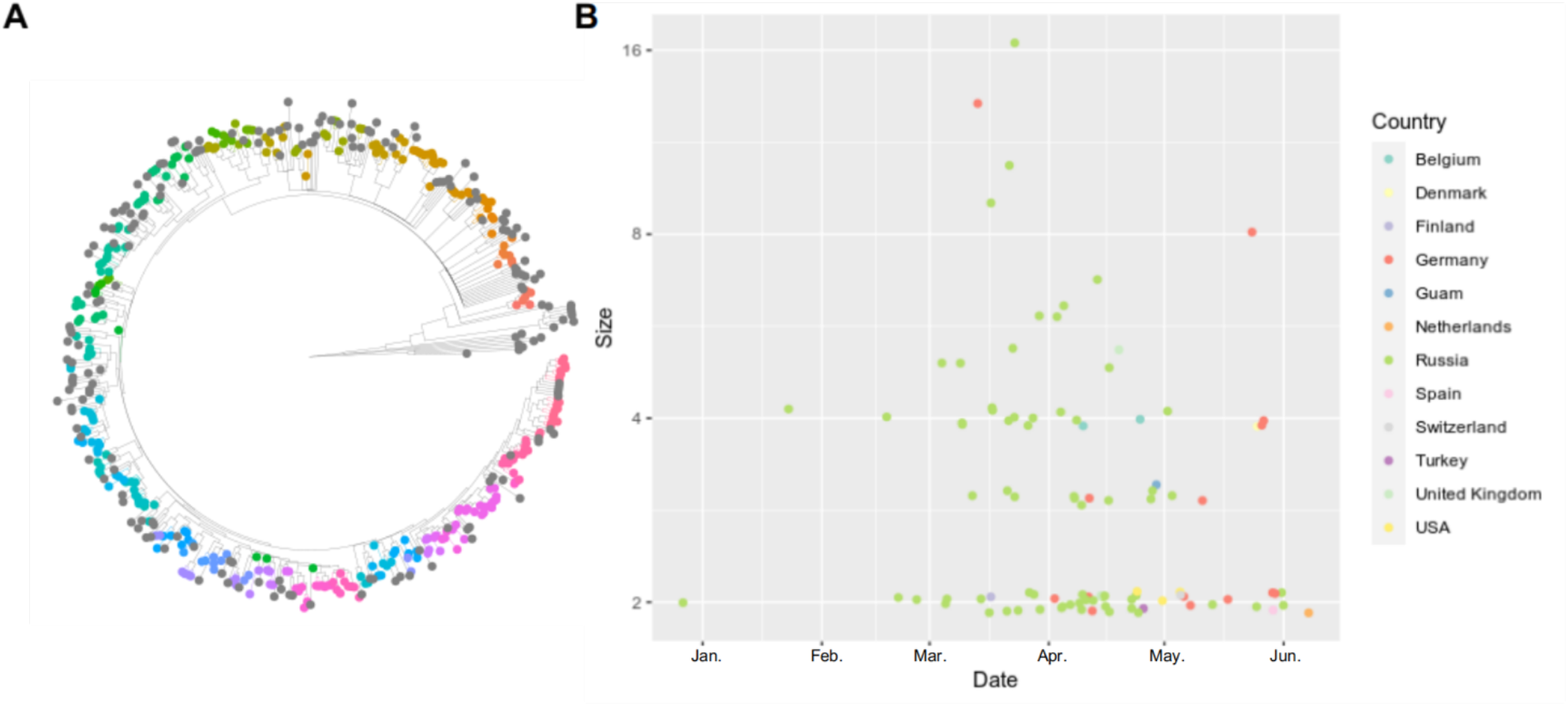
B.1.1.523 Transmission clusters identified using the phylogeny tree. Sequences with identity larger or equal to 99.3 % to the Latvian B.1.1.523 sequence (EPI_ISL_1590462) were added to the analysis. Colours indicate sequences belonging to the same transmission cluster. Grey colour marks sequences that were not assigned to any transmission colour (A). Colour encodes the country of the most recent common ancestor (MRCA) of all sequences that constitute a cluster. Y axis represents size of a cluster and Y axis denotes the date inferred for the MRCA sequence (B).

Most of the cases transmission origin country (country inferred for a cluster MRCA sequence) and the target country (country of a sequence indicated in the GISAID metadata) was Russia and as we see in the Figure 6B majority of the transmission events happened within Russia. Evidently The largest number of cases were transmission origin and target countries were different resembles transmission from Russia to Germany. The data indicate one reverse type (Germany to Russia) transmission. As of the date of writing this article, Germany can be considered as a reservoir of the B.1.1.523 lineage. As for May 2021 the B.1.1.523 and Delta variants represented 0.002% and 0.054% respectively, while the data of August 2021 shows a steady increase in detected cases to 0.327% and 32.358% respectively. The overall number of sequenced cases in Germany is becoming overwhelmed by Delta variant, however the lineage B.1.1.523 lineage seems not to be fading away (Fig. 8). Data shows that B.1.1.523 is able to steadily spread without interference with the Delta variants and can be considered as a predominant background lineage of SARS-CoV-2. As indicated in the Figure 6. the spread of the lineage (based on GISAID submissions) in Russia diminished while it remains evidently circulating in Germany. It is unclear whether B.1.1.523 is completely out of circulation in Russia or its frequency dropped below the sensitivity threshold of SARS-CoV-2 genomic surveillance in this country.

**Figure 8.**
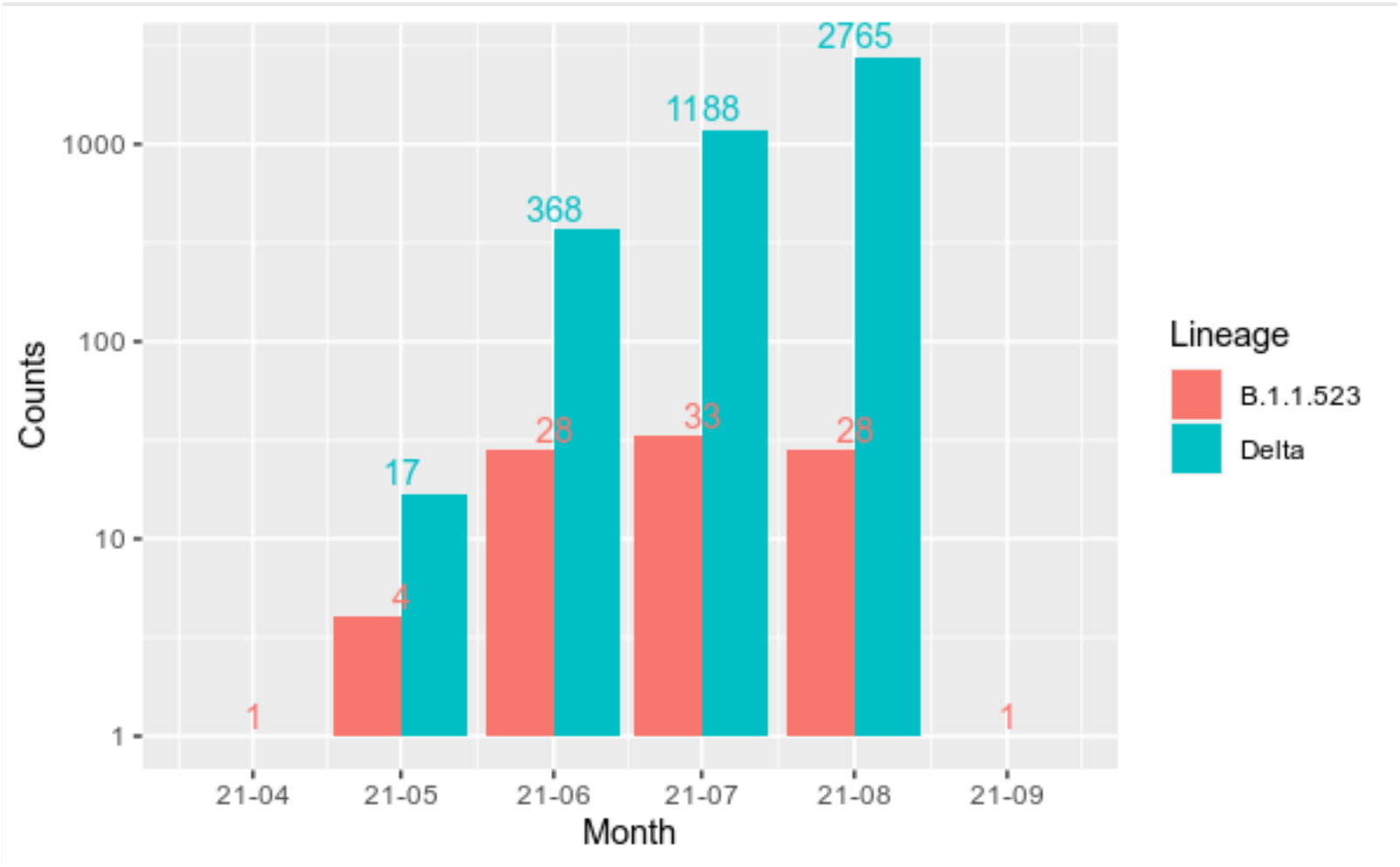
Newly sequenced cases of B.1.1.523 and Delta lineages in Germany. Data number of new cases per month.is based on GISAID metadata (August 31st, 2021). Numbers indicate the total number of sequences deposited in GISAID.

### B.1.1.523 antibody escape

In Figure 9A the modelled free energy of complex separation is given as calculated for the three sequence variants considering the NTD-antibody complex. As Rosetta documentation states^11^ ΔG of complex separation (dG_separated) shows the change in Rosetta energy when the interface forming chains are separated, versus when they are complexed - therefore the lower the value the more energetically favourable is complex separation and less favourable is complex formation. In other words - the higher the value - the less likely should be the neutralization by the antibody. Pairwise comparisons using Wilcoxon rank sum test with Benjamini-Hochberg correction for multiple comparisons indicated that all three cases significantly differ from each other (p values are given in Fig. 9A upper right corner).

**Figure 9.**
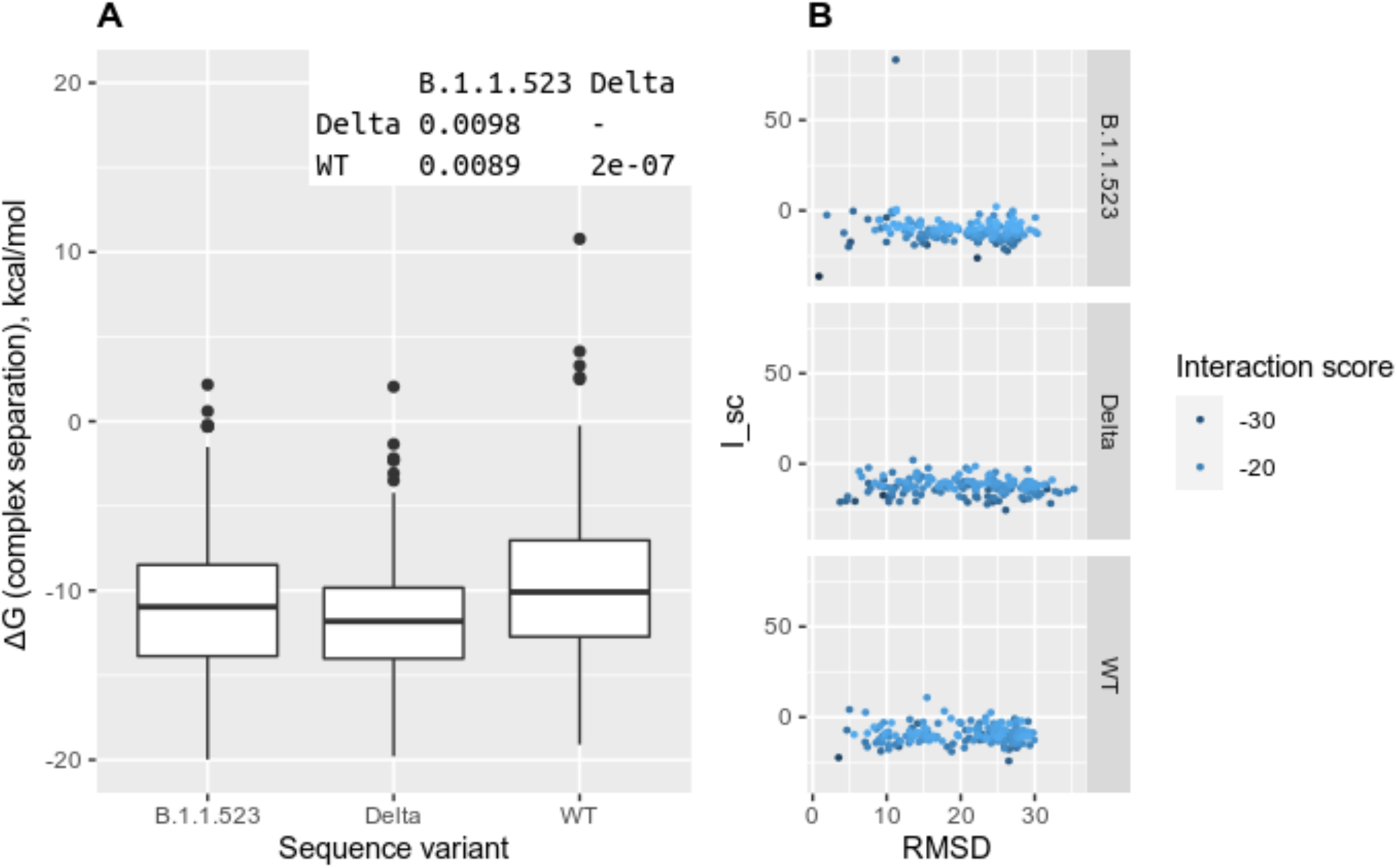
Escape effects of the d156_158 mutation (B.1.1.523) and del156_157&R158G mutations (delta variant) based on NTD-directed neutralizing antibody 4-8 Fab (PDBID: 7LQV). Predicted complex separation ΔG values for the mutants and the wild type complexes (A). The relationship between structure deviation from the starting structure used for the docking with SnugDock and the I_sc score (B).

All calculated FoldX binding energies are given in the supplementary file FileS1.xlsx. At least in four cases a significant synergy in the effect of the E484 and S494P mutations were observed: notably for the antibodies H11-H4_6ZH9, H11-D4_6YZ5, Sb45_7KGJ, Sb16_7KGK (Fig 10A). For the most part the most significant antibody escape effect was noticed by the E484K mutation; however, in some cases the S494P effect was also prominent. The largest effect by the S494P mutation was observed for the E_7KN5^12^ antibody structure (Fig. 10B): the S494P mutation increased the binding ΔΔ G by 74 % and the additional effect of E484 mutation was negligible (Fig. 10A).

**Figure 10.**
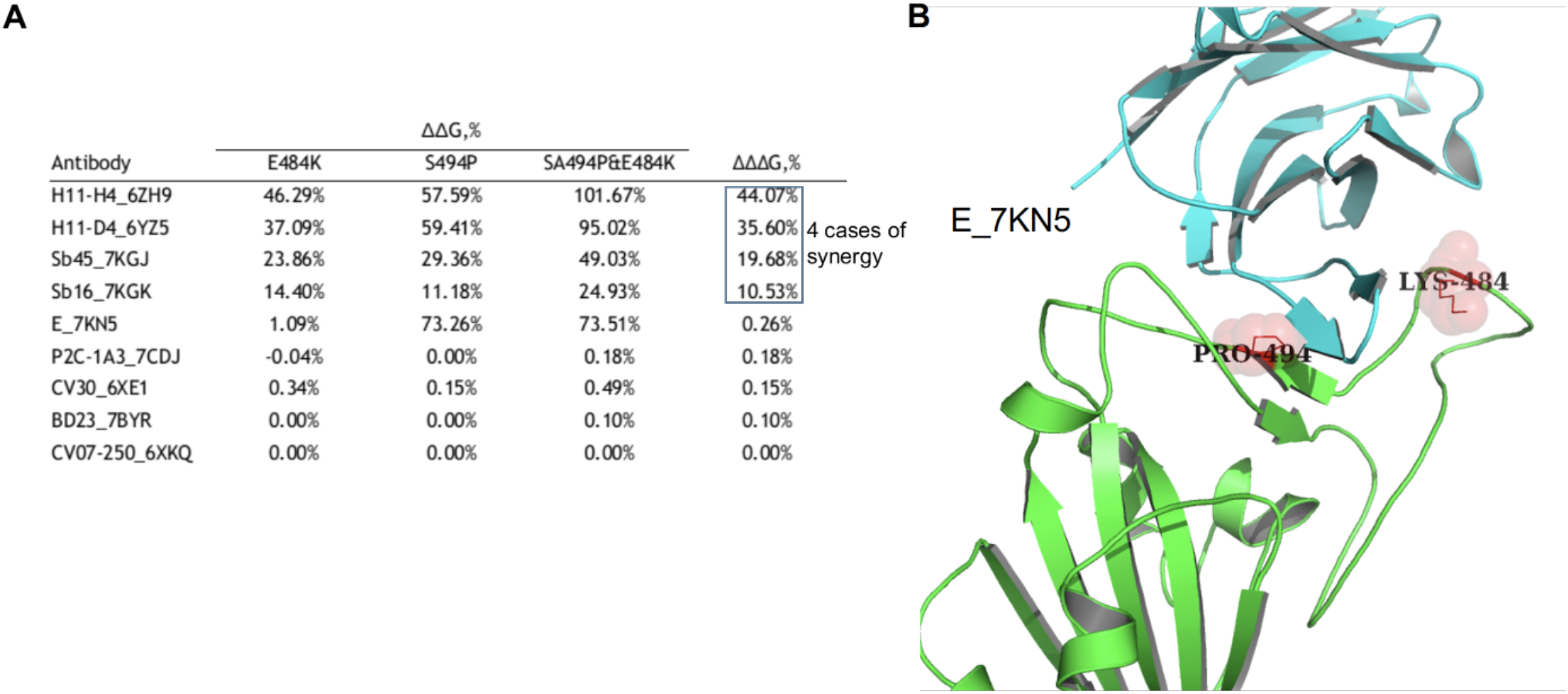
Escape effects of the E484, S494P mutations and their combination. The ΔΔG values indicates relative increase in binding energy compared to the wild type structure as inferred from the FoldX calculations. The ΔΔΔG indicates the minimum difference between the ΔΔG of the double mutation and any of the two single point mutations. The large the value - the large the synergy (A). The structure of the antibody and receptor binding domain of the S protein complex which was affected by the S494P mutation most significantly (PDB ID: 7KN5) (B).

## DISCUSSION

We have identified a new SARS-CoV-2 virus lineage with multiple mutations associated with immune escape and reported this to Pango at 5’th of May [https://github.com/cov-lineages/pango-designation/issues/69], which mandates the new lineage designation B.1.1.523. This Lineage was first determined in March 2021 and at the time of writing this article, the total amount of cases has reached 598 over 32 countries 12. It is likely that the rapid increase in circulation of Delta variant could have diminished the rise of B.1.1.523 lineage, however, the spread of the novel SARS-CoV-2 lineage not only has not ceased, but even has started to rise.

Currently, a vast growth of B.1.1.523 can now be observed in Germany. Interestingly, the transmission of this lineage has diminished in Russia, where it was most expected to rise. This can be explained by different diagnostic strategy approaches in Russian Federation, where the testing is performed on non-randomly selected sources in the country. Alternatively, this could be explained by the steep rise of Delta variant in Russia, which started a month earlier than in Europe. E.g. in mid June we had >80% delta, while in Germany the same frequency of Delta was observed only in mid July.

The B.1.1.523 lineage possesses three or more mutations that characterizes SARS-CoV-2 VOCs, including S:D156-158 deletion, S:E484K and S:S494P. D156-158 deletion at β-hairpin antigenic supersite, that is located at the same region typical for the Delta variant (E156G and 157-158del)^13^. E484K mutation has been detected in Beta variant (B.1.351) and VUM Zeta (B.1.1.28). The mutation is in the genomic region coding SARS-CoV-2 spike protein, and it appears to have a significant impact on the body’s immune response and possibly, vaccine efficacy. On February 1^st^, Public Health England (PHE) announced that the Covid-19 Genomics (COG-UK) consortium had identified this same E484K mutation in 11 samples carrying the UK variant B.1.1.7 (sometimes called the Kent variant), after analysing 214 159 sequences^14.^

An in vitro study for SARS-CoV-2 spike protein mutations that are responsible for antibody evasiveness has identified that S494P mutation 15 reduce SARS-CoV-2 neutralization by 3-5-fold in some convalescent sera. However, this mutation was not as potent at neutralization as E484K 16. The results show that S494P mutation increases the spike protein stability. Also, applying docking by HADDOCK displayed higher binding affinity to hACE2 for mutant spike than wild type possibly due to the increased β-strand and turn secondary structures which increases surface accessibly surface area (SASA) and chance of interaction. Currently deposited sequences in GISAID do not support hypothesis that S:156_158del were combined with S:E484K and S:S494P during a recombination event; rather it looks like that initially the triple deletion was introduced and then followed addition of S:E484K and S:S494P.

We have showed by molecular modelling that in at least one case of antibody the triple deletion del156-158 could decrease interaction. The combination with other immune escape enhancing mutation at RBD this could result in highly resistant variant to immunity that was formed by the initial virus variants. Delta variant also poses sequence changes at the S protein residues 156-158 that can induce immune escape and recombination with the B.1.1.523 variant or de novo introduction of the N484K and S494P mutations could make the Delta variant even more dangerous. The case with Turkey sequences is controversial. The sequences at the S region significantly deviated fragments with many “private” mutations. These sequences have been seen across several submissions; hence, these might be not artificial. If these sequences are not artifacts, then the Turkey sequences classified by Pangolin as belonging to B.1.1.523 lineage, resulted after a recombination event between a highly diverged Turkey variant and a typical B.1.1.523 lineage sequence. Turkey has been an extensive place of the virus spread with limited control and reportability ^17^; therefore, highly diverged variants could have evolved.

The results indicate that Pangolin classification should not be taken with granted. Out of the two Turkey sequences that were assigned to the B.1.1.523 lineage, only one had characteristic to the lineage SNP’s at the S protein, the other one (Turkey/HSGM-B11931/2021) has mutations characteristic for the Delta variant (double deletion and G residue at 156-158 region).

## CONCLUSIONS

Presence and spread of SARS-CoV-2 B.1.1.523 lineage is evident regardless of the rapid spread of the delta variant. This variant needs to be carefully observed and studied to keep a look out for new mutations, that may cause even more harm in the Covid-19 pandemic. It is also important to monitor other SARS-CoV-2 variants to keep track if this or similar mutations occur spontaneously or by recombination.

## METHODS

### Collection of SARS-CoV-2 sequences and initial data processing

Sequences used for the analyses were downloaded from GISAID^18^ as for date of August 31^st^, 2021. Fasta files and metadata were extracted using ncov-ingest tool [https://github.com/nextstrain/ncov-ingest (cloned at 2021 04 19)]. Lineages for all downloaded sequences were assigned using pangolin 3.1.11 (pangoLEARN 2021-08-24 and pango-designation v1.2.66)^19^ [https://github.com/cov-lineages/pangolin].

General sequence quality evaluation, extraction and alignment of S protein sequences, variant calling was performed using Nextclade 1.3.0^20^ [https://github.com/nextstrain/nextclade].

### Transmission cluster analysis

In order to elucidate potential origin of the lineage and transmission cluster, a phylogeny analysis of full genomes representing a small subset of GISAID has been done. The chosen sequences for analysis composed from the union of two sets of sequences: (i) sequences that were assigned B.1.1.523 lineage by pangolin, (ii) sequences that were at least 99.3 % identical to the Latvian B.1.1.523 sequence EPI_ISL_1590462 and number of matched residues makes up equal or more than 95% of the reference sequence. The reference sequence was chosen as it was closest to the one of the first this lineage sequences sequenced at Lithuania but with smaller gaps regions. The alignment against the GISAID sequences was conducted using minimap2 2.20-r1061 [https://github.com/lh3/minimap2/]. The limits were chosen arbitrary after several tries looking for cut-offs resulting in a set of sequences that includes majority of sequences from the lineage and some more diverged ones that are classified as belonging to other lineages by pangolin. The sequences with quality control overall status being bad or having more than 1000 bps missing (as indicated by Nextclade analysis) were discarded. The maximum likelihood tree was calculated using a modified version of Nextstrain workflow [https://github.com/nextstrain/zika (cloned at 2021 05 01)]. The tree was build using IQ-TREE with 2.1.2 General time reversible model with unequal rates and unequal base frequencies were used^21^ allowing for a proportion of invariable sites together with discrete Gamma model^22^. Ultrafast bootstrap with 1000 replicates was used. The maximum likelihood emergence time and origin of country for inner nodes were calculated by treetime 0.8.1 [https://libraries.io/pypi/phylo-treetime] as described by the aforementioned workflow. The set of sequences collected as described above were clustered into transmission clusters using Phydelity v2.0 [https://github.com/alvinxhan/Phydelity]^23^.

### S protein phylogeny analysis

The S protein-based phylogeny was based on the S protein sequences extracted from GISAID by the Nextclade and aligned to the reference COVID-19 sequence. Sequences shorter than 1175 residues or having more than one stop codon or having any number of undetermined residues were discarded. Sequences were further clustered into identical sequence clusters using CD-HIT v4.8.1 (command line option “-c 1.0 “. The sequences representing all high-quality S protein variants were used for maximum likelihood tree calculation by VeryFastTree 3.0.1 [DOI: 10.1093/bioinformatics/btaa582] using LG substitution model. The sequence alignment that was used to construct the tree was composed from the alignment produced by the Nextclade leaving only the representative sequences of the clusters and CAT approximation with 20 rate categories. lg -gamma”. Also command line flags that should increase calculations accuracy were added: “-spr 4 -mlacc 2 -slownni -double-precision “. The tree was re-rooted using sequence matching to the EPI_ISL_402124 as an out-group using gotree 0.4.1 [https://anaconda.org/bioconda/gotree/files].

Ancestral states for inner nodes for 156, 157, 158, 484, 494 positions of protein S were inferred using command line version GRASP-suite^25^. This tool for inference was chosen due to high speed and, most importantly, ability to handle insertion and deletions. The same substitution model that used for phylogeny tree was also used in this case (LG). The resulting tree was analysed detected potential changes in the haplotypes was done using a custom script written in julia 1.6 exploiting capabilities of the NewickTree library [https://github.com/arzwa/NewickTree.jl]. The tree was trimmed to keep only those inner nodes that leads to leaves containing the triple deletion at 146-148 positions and either E484K, S494P and visualised using ggtree 3.0.426. Additionally, a set of full genome sequences has been composed matching the leaves of the aforementioned trimmed tree and corresponding maximum likelihood tree was calculated using the aforementioned Nextstrain workflow.

### Analysis of potential recombination events

The focused set of sequences was composed based on the sequences used for the S protein phylogeny. The set further narrowed to the sequences containing either the triple deletion at 156-158 positions or a combination of E484K and S494P. The detection of recombination events at DNA level was done using PoSeiDon workflow27 [https://github.com/hoelzer/poseidon cloned at 2021 09 28] that runs GARD program to identify recombination events^28^. The genomic regions matching to the S protein were used. As the workflow is limited to 100 sequences the initial set 110 was clustered to 100 sequences using cd-hit-est program from the CD-HIT package 4.8.1 using identity cut-off equal to 0.9998658. The detection of recombination effects at protein level was done using detREC tool29 [https://github.com/qianfeng2/detREC_program (cloned at 2021-10-01)]. The recombination detection was conducted using either genomic or protein sequence corresponding to the S protein. The full genome scale recombination analysis was done for the same set of 110 sequences using 3SEQ program^28.^

### Antibody escape effect estimation

Two S protein sequence variants 156_158del (B.1.1.523 lineage) and 156_157del & R158G (delta variant) were modelled using Rosetta package (2021.16.61629_bundle)^31^. The escape effect was evaluated based on the structure PDBID: 7LQV containing a NTD domain targeting binding antibody. The S protein structure from the “A” chain with matching antibody was used. The S protein residues from 283 position to the “N” terminus were discarded. The PDB structures for fragment library generation and other required databases for Rosetta were downloaded at 2021 06 02. The S structure models were created using a comparative modelling approach. The antibody structure including side chains was kept constrained using CoordinateConstraintGenerator. For the relaxation part “InterfaceRelax2019” script was used. The lowest energy model was chosen from 200 models. The antibody docking was done using the SnugDock program^32^ from the Rosetta package, using these non-default flags: “-auto_generate_h3_kink_constraint - h3_loop_csts_hr -h3_filter false - docking_centroid_inner_cycles=20 -docking_centroid_outer_cycles=2 “. for each model docking was run 300 times and the top 175 structures based on the interface score were chosen for analysis.

Antibody escape effect of E484K and S494P mutations were analysed using FoldX 5 program^33^. The antibody structures targeting the receptor binding domain of the protein S were downloaded from the CoV3D^34^ as available at 2021 05 01. The mutations into the complexes were introduced by consecutively applying FoldX command: RepairPDB, BuildModel, AnalyseComplex. The residues at 484 and 494 positions were modelled either to the residue matching the mutation or to the wild type residue and changes in free energy upon binding were evaluated.

## Data Availability

All data produced in the present work are contained in the manuscript

